# No Detectable Surge in SARS-CoV-2 Transmission due to the April 7, 2020 Wisconsin Election

**DOI:** 10.1101/2020.04.24.20078345

**Authors:** Kathy Leung, Joseph T. Wu, Kuang Xu, Lawrence M. Wein

## Abstract

We analyze confirmed cases and new hospitalizations in Wisconsin in the weeks surrounding the April 7, 2020 election, and find no evidence of a surge in SARS-CoV-2 transmission.

---

The April 7, 2020 Wisconsin election produced a large natural experiment to help understand the transmission risks of the severe acute respiratory syndrome coronavirus 2 (SARS-CoV-2). Up to 300,000 people voted in person [1-2] and waiting times in Milwaukee averaged 1.5-2 hr [3]. Poll workers had surgical masks and latex gloves, hand sanitizer was made available to voters, isopropyl alcohol wipes were used to clean voting equipment, and painting tape and signs were used to facilitate social distancing [2].

Wisconsin tracks cases confirmed by testing (Fig. 1A) and throughout April 2020 have restricted testing to frontline workers and those hospitalized with serious illness [4]. We used a deconvolution-based method to reconstruct the SARS-CoV-2 epidemic curve by dates of infections rather than dates of reporting by health authorities, and then used two different methods [5]-[6] to estimate the instantaneous reproduction number *R*_*t*_, which is the average number of secondary cases generated by one primary case with the time of infection on day *t*, from March 25 (the start of the safer-at-home order) through April 18 (see the Supplementary Appendix).

**Figure 1.**
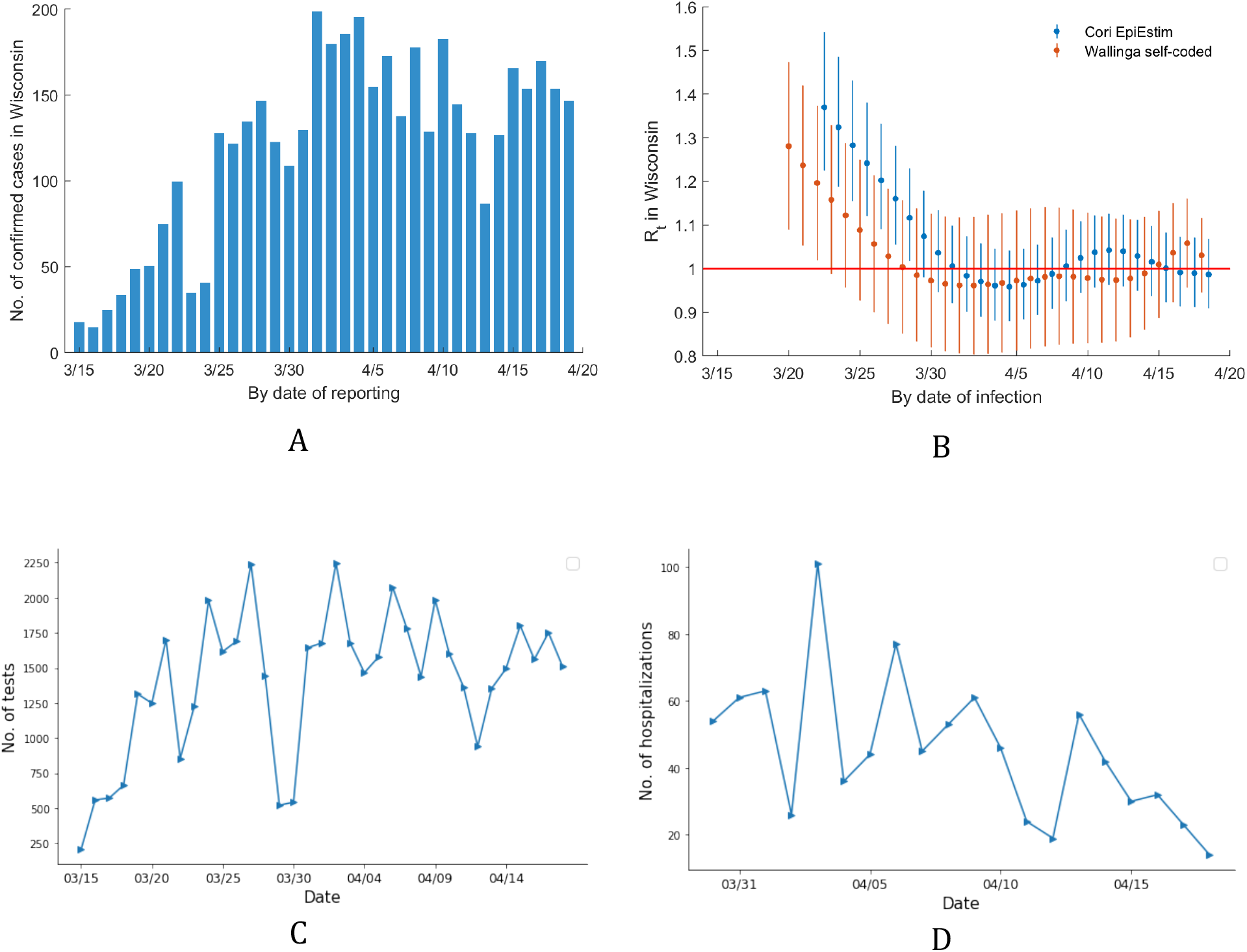
SARS-CoV-2 Dynamics Surrounding the April 7, 2020 Election in Wisconsin. Panel A shows the number of daily confirmed SARS-CoV2 cases in Wisconsin from March 15 to April 19. Panel B shows the estimated instantaneous reproduction number *R*_*t*_ (along with 95% confidence intervals) each day from March 25 (the start of the safer-at-home-order in Wisconsin) to April 18 using two different methods. Panel C shows the number of SARS-CoV-2 tests performed each day from March 15 to April 18. Panel D shows the number of new SARS-CoV-2 hospitalizations in Wisconsin on each day from March 30 to April 18. In generating the curve in Panel C, a possible mis-entry in the original data set [4] led to the cumulative test count on March 29 being smaller than the day prior; in response, we replaced the March 29 cumulative case count by the average value between March 28 and 30.

As seen in Fig. 1B, there is no detectable spike in *R*_*t*_ on April 7. The number of SARS-CoV-2 tests performed in Wisconsin has been relatively stable throughout April [7] (Fig. 1C), suggesting that reduced testing capacity in the days after April 7, which could have censored some of the April 7 infections, did not occur. Moreover, new SARS-CoV-2 hospitalizations in Wisconsin have steadily declined throughout April (Fig. 1D), from a high of 101 on April 3 to a low of 14 on April 18 [7], suggesting that daily new hospitalizations are much less than testing capacity.

The lengths of the incubation period and the reporting delay imply that April 7 infections would not be reported until April 17 on average, with most cases being reported during April 11-22. Taken together, there is no evidence to date that there was a surge of infections due to the April 7, 2020 election in Wisconsin, which has a relatively low level of SARS-CoV-2 transmission in the US.

Finally, the Wisconsin Department of Health Services announced on April 22 that 19 people who either voted in person or worked at the polls on April 7 have tested positive for SARS-CoV-2, although several of these people also experienced non-voting exposures [8]. This fact is not inconsistent with our population-level analysis, because 19 cases is small relative to the total number of confirmed cases in Wisconsin. To put this information into perspective, if we assume that the SARS-CoV-2 fatality rate among symptomatic patients who were physically capable of voting in person on April 7 (e.g., not including nursing home residents) is 1% (using the fatality rate of known cases for people aged <60 [9]), then we would expect 0.19 deaths out of 300,000 people, which is the fatality risk of driving an automobile approximately 50 miles [10]. However, in addition to the individual risk of voting on April 7, there is the community risk: how many downstream cases will these 19 original cases generate? According to Fig. 1B, the reproduction number in Wisconsin has been hovering near the value of one for all of April. If this value was much larger than one (as it was in, say, January) then these 19 cases would cause a lot of downstream damage, and if this value was clearly smaller than one then they would cause minimal damage. But a value near one, coupled with the small number of cases, means that it is very difficult to reliably predict the amount of downstream damage.

Taken together, it appears that voting in Wisconsin on April 7 was a low-risk activity.

## Data Availability

All data are easily available from the references in the manuscript, and are plotted in Figure 1.

## SUPPLEMENTARY APPENDIX

The instantaneous reproductive number *R*_*t*_ was defined as the average number of secondary cases generated by one primary case with the time of infection on day *t*. If *R*_*t*_ > 1 the epidemic is expanding at time *t*, whereas *R*_*t*_ < 1 indicates that the epidemic size is shrinking at time *t*.

Since the epidemic curve of Wisconsin is based on the dates of test confirmation, we use a deconvolution-based method to reconstruct the SARS-CoV-2 epidemic curve by dates of infection [1-2]. Let *f*_*incubation*_ be the probability density function (pdf) of the incubation period, and *f*_*onset confirmation*_ be the pdf of the time between symptom onset and test confirmation. We assume *f*_*incubation*_ and *f*_*onset confirmation*_ are independent such that the pdf of the time between infection and confirmation is

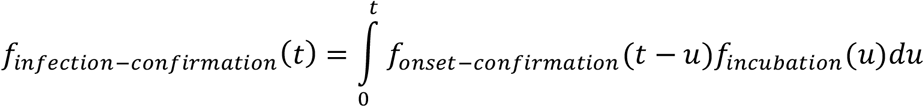

We use *f*_*infection-confirmation*_ to deconvolute the time series of the daily number of confirmed cases to reconstruct an epidemic curve of daily number of new infections. We assume the incubation period distribution is gamma with mean and SD of 5.2 and 2.3 days [3]. We assume that the distribution of the time between symptom onset and confirmation is gamma with mean and standard deviation (SD) of 4.3 and 3.2 days, based on 186 cases reported in Jan-Feb 2020 in Beijing [4]. With the epidemic curve by dates of infection in hand, we applied two different methods -- developed by Wallinga and Teunis [5] and by Cori et al. [6] -- to estimate *R*_*t*_ using the R package EpiEstim. We assume the generation time distribution is approximately the same as the serial interval distribution, which was inferred to be gamma with mean 5.4 and SD 4.7 days from the dates of symptom onset of 56 infector-infectee pairs from mainland China [4].

## Notes

### Competing Interest Statement

The authors have declared no competing interest.

### Funding Statement

No funding

